# ‘The Recommended Deltoid Intramuscular Injection Sites in the Adult Population: A Cadaveric study with an Orthopedic Perspective’

**DOI:** 10.1101/2024.06.14.24308929

**Authors:** Sundip Hemant Charmode, Abhishek Kumar Mishra, Simmi Mehra

**Affiliations:** Department of Anatomy, All India Institute of Medical Sciences, Rajkot, Gujarat; Department of Orthopedics, All India Institute of Medical Sciences, Rajkot, Gujarat

**Author notes:** **Corresponding author:** Name: Dr. (Lt. Col.) Abhishek Kumar Mishra, Institution: All India Institute of Medical Sciences, Rajkot, Department : Orthopedics, Address: Department of Orthopedics, AIIMS Rajkot, Gujarat-360110,; 9633239685. **Dissemination history (where applicable) :** This article has not been presented anywhere or the content of this article has not been published anywhere before. Section of the journal to which the article is addressed : Original Article. **Author contributions:** Dr. Sundip Charmode initiated the idea, designed, and drafted the article. Dr. Lt. Col. Abhishek Kumar Mishra contributed to data acquisition, analysis, and interpretation of acquired data. Dr. Simmi Mehra reviewed the manuscript and gave valuable suggestions.

**Keywords:** Deltoid intramuscular injection, Periosteal injury, Peripheral nerve injury, Clinical Trail, Cadaveric study

## Abstract

**Background:** The deltoid is a common site for intramuscular injections, but guidelines for administration lack standardization. Global researchers propose various techniques, and recent study reports indicate a 1.5-15% incidence of nerve palsies due to injections. Our cadaveric study is aimed to standardize the deltoid intramuscular injection sites in the Southeast-Asian population.

**Methods:** A cadaveric study of a 2-year duration was conducted in the Department of Anatomy in which twelve upper extremity specimens were dissected by the end of the pilot phase. Anthropometric measurements of deltoid muscle along with the distance of underlying neuro-vascular structures like the Axillary nerve and Posterior Circumflex Humeral Artery were measured from neighboring bony landmarks.

**Results:** In adults, in anatomical position, the mean distances of the Axillary nerve, and Posterior Circumflex Humeral Artery from the mid-acromial point are 8.19 ± 0.616 cm; and 8.66 ± 0.968 cm respectively. The deltoid thickness at 3, 5, and 7 cm from mid-acromial point was observed to be 1.079 ± 0.13 cm (0.5 cm to 1.78 cm), 1.599 ± 0.12 cm (1 cm to 2.96 cm), 1.815 ± 1.0 cm (1.2 cm to 2.5 cm) respectively. The acquired qualitative and quantitative data were tabulated, graphically represented, and statistically analyzed.

**Conclusions:** The deltoid IMI must be given at or below the level of the midpoint of the deltoid muscle, but never in the upper half. We recommend a site, 4 fingerbreadths / 9cm below the mid-acromion point as the safest site to avoid injury to any underlying neurovascular structures.

## Introduction

Globally, the deltoid is the preferred IM site in clinical practice [1]. Many other IM sites have been considered over the deltoid based on the risk of injury to the underlying vessels and nerves. However, a paucity of uniform guidelines and faculty advisories persists for IM administration by healthcare professionals [2]. The incidence of intramuscular injection-related peripheral nerve injuries ranges from 1.5% to as high as 15% [3]. These observations were confirmed by another study conducted at PD Hinduja hospital and Seth GS medical college Mumbai in 2019 by Desai et. al. on 354 patients where incidence of IM induced nerve injuries was observed as 82.5% [4]. More than 50 % of deltoid intramuscular injections (IM) are given indiscriminately by untrained staff in unregistered settings in clinical practice, especially in Southeast-Asian countries [3,5]. In the East-Asian and West, extensive research is done namely Cheung K et al., (2009) USA [6]; Chen YF et al., (2012) China [7]; Ye-Gyung Kim et al., (2022) Korea [8]; and Nakajima et al., (2017) Japan [9]. In India, only two research studies namely Patra A, et al., (2018) [10] and Gurushantappa PK, (2015) [11] exists on the anthropometry of axillary nerve. In a recent clinical trial conducted by Surraj S, Chandrupatla M, and Kusneniwar GN. in south Indian population, two sites first (one-two centimeters above the deltoid insertion) and second (midway between the middle of the arm and the deltoid insertion) were reported to be safe for deltoid intramuscular injection [12]. As per our theories and experiences, we contradict the above-mentioned sites, as we fear injury to periosteum of humerus at any site close to the deltoid insertion. In addition to this, a systematic review was conducted by the authors in 2022 that confirmed few deficiencies in IM guidelines issued by regulatory bodies of India and across the world [13]. This cadaveric study is targeted to validate our review findings thereby aiming to propose a safe site for deltoid IM with the least chance of injury to the underlying neurovascular structures by conducting the anthropometric measurements of the deltoid muscle and determining the sites of various neuro-vascular structures underlying or related to the deltoid muscle.

## Methods

### Study design and population

A cross-sectional study of observational type was conducted in the Department of Anatomy, in collaboration with the department of orthopedics. The study duration was of 2 years after approval from Institutional Ethical Committee of the institute. Clinical Trial Registry India (CTRI) registration could not be done as cadaveric studies are not included in CTRI registry. A sample size of 73 cadavers (146 specimens) was calculated using the prevalence rate of symptomatic axillary nerve compression as 5% based on the reference study, Surraj S et al. 2022, that was conducted at the Department of Anatomy of AIIMS Bibinagar [12].

### Sample size calculation

For the sample size calculation, the prevalence rate of symptomatic axillary nerve compression was taken as 5%. This was taken from the reference study, Surraj S et al. 2022, AIIMS Bibinagar [12]. The formula to calculate the sample size in medical studies from prevalence rate was proposed by Mohammad Amin et al. 2012 Iran [14].

Considering ‘n’ as the sample size for the study group, P was 5, Q was 95, L (precision) was taken as 5% with a 95% Confidence Interval and the power of the study was considered as 80.0. Using the formula, sample size (n) = Z x PQ/L, n was calculated as 72.6 which was rounded off to 73. So, sample size (n) amounted to 73 cadavers. Looking at the cadaver availability in the department, it was decided to conduct a pilot study in the first year of the project on the available 6 cadavers. The pilot study findings shall be validated in the second year of the project.

### Eligibility criteria

Eligibility criteria (Inclusion and exclusion) for the cadavers were framed by the investigators. The inclusion criteria for cadavers were i) Embalmed human bodies using standard embalming techniques, ii) Specimens not displaying (or without history of) any congenital/acquired deformity or metabolic disorders or malignancy, or any other debilitating disease. iii) Specimens without history of/not displaying any fracture of humerus, scapula or pectoral girdle. iv) Specimens without history of any surgery performed on humerus, scapula or pectoral girdle. v) Specimens not displaying any scar, injury or deep wound on the arm and shoulder joint region. The exclusion criteria for cadavers were i) Un-embalmed human bodies, ii) Specimens displaying (or with history of) any congenital/acquired deformity or metabolic disorders or malignancy, or any other debilitating disease, iii) Specimens with history of/displaying any fracture of humerus, scapula or pectoral girdle, iv) Specimens with history of any surgery performed on humerus, scapula or pectoral girdle, v) Specimens displaying any scar, injury or deep wound on the arm and shoulder joint region.

### Ethical approval

This study was conducted as phase 2 of an Intramural research project [Funded] in the Department of Anatomy in collaboration with the Department of Orthopedics of the institute. The project was submitted and presented at the Research Review Board (RRB) and after its approval, it was forwarded to the Institutional Ethical Committee of the institute. The IEC approval was received on 13^th^ September 2023 with the PROTOCOL ID: F-IM/15/2023. The approval letter ref no. O.W.No./AIIMS. RKT/IEC/54/2023 dated 13^th^ September 2023.

### Clinical Trial Registry India (CTRI) registration

After the IEC approval, the project proposal was submitted to the Clinical Trial Registry of India (CTRI) and the registration was returned on 27^th^ September 2023. The reference number is REF/2023/09/072948 and the link is https://www.ctri.nic.in/.

### Data collection form

To collect the general information about the cadaver a specially designed, data collection form (in English) was formulated to collect demographic data like age, gender, religion, occupation, education, family contact details, postal address, cause of death, medical history, surgical history of concerned part, diagnosis, mode of treatment received (conservative or surgical) for the condition, unilateral or bilateral involvement, and duration of the illness. The medical records were referred in order to collect this information. In event of the information unavailable in the medical case records, the family members were contacted to extract this information. In addition to this, the anthropometry findings of the concerned body structure in the cadavers were recorded. Informed consent proforma was not required in this study as all the study participants i.e. the cadavers were voluntarily donated to the institute.

### Data collection procedure

Standard methods were used to dissect the concerned body region that included the cautious removal of skin along with subcutaneous tissue from the shoulder and arm region while retaining the fascia over the deltoid muscle. In this study, we focused on the acromial origin of the deltoid muscle (DM). The anthropometric measurements of the DM were done using digital vernier calipers and anthropometric tape. Each measurement was repeated to an accuracy of within 1 mm, and the average of both measurements, rounded up to 1 mm, was accepted as the final result. Two investigators conducted the measurements separately on each cadaver at different times in the day. A research study piloted by Ye-Gyung Kim et al. (2022) in Korean population was referred to as reference study for the landmarks used for conducting anthropometric measurements in the present study [8]. Before taking the measurements, bony landmarks were identified and marked using specimen marking pins on the concerned region namely the anterior point of the lateral border of acromion (AP), the posterior point of the lateral border of acromion (PP), and the acromion. The origin point (OP) was defined as the midpoint of a line connecting the AP and PP at the acromion level and a vertical line was constructed by connecting the OP to the lowest point of the deltoid tuberosity (DP) as shown in Figure 1.

**Figure 1:**
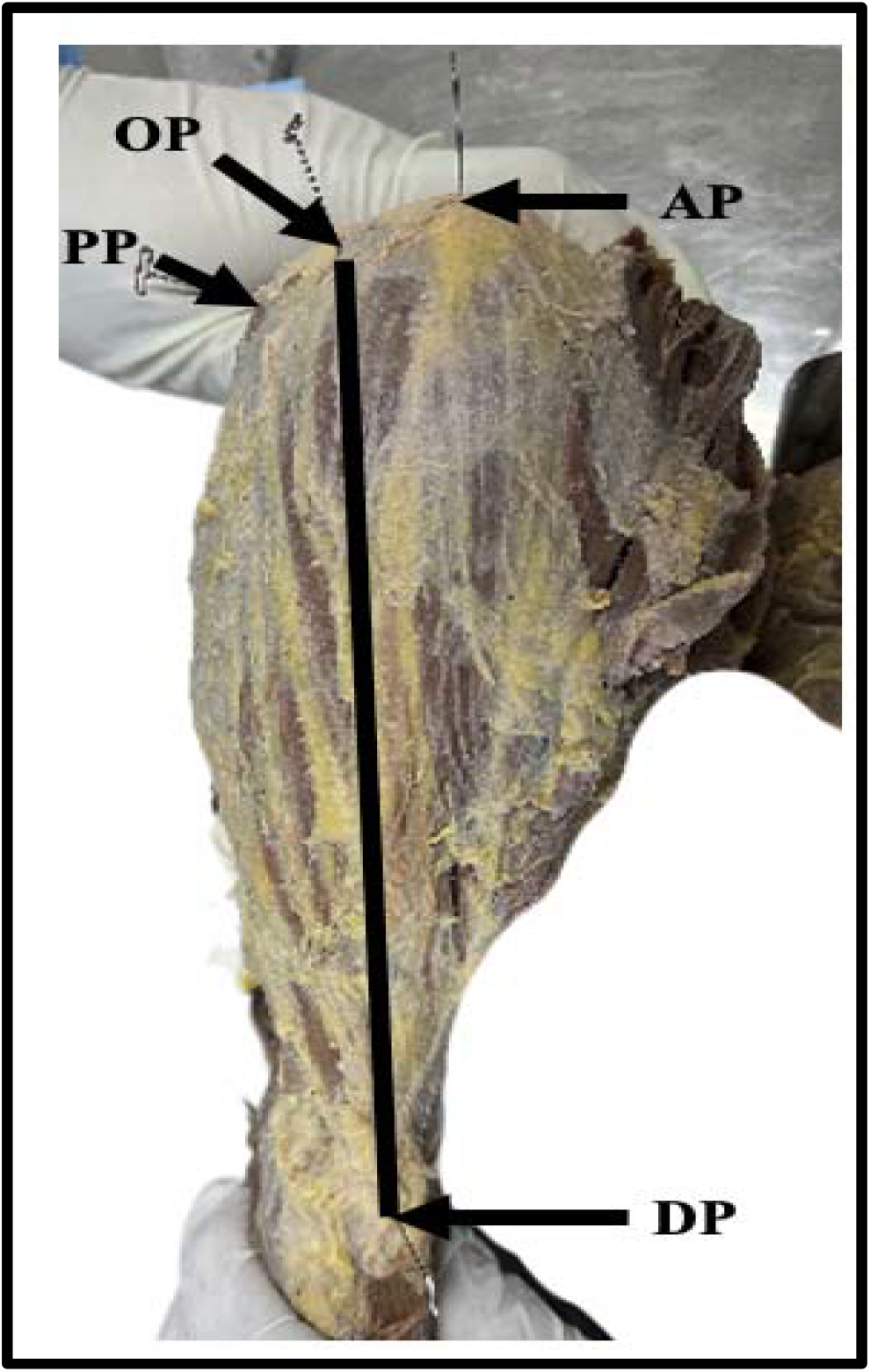
Bony landmarks for reference line measurements. Courtesy: This image is obtained from the Department of Anatomy, AIIMS Rajkot, Gujarat, India.

The thickness of the deltoid muscle, skin, and soft tissue was measured using digital calipers (resolution 0.01mm/0.0005, In size 1108-150: Digital caliper, range 0-150 mm, Accuracy +/- 0.03 mm) at 3, 5, and 7 cm distance from the OP. The length of deltoid was measured from (OP) to (DP). The distance of the Anterior Branch of the Axillary Nerve (AXN), and Posterior Circumflex Humeral Artery (PCHA) from the mid-acromial point were measured.

### Data categorization

The data collected from the cadavers was categorized into reference line measurements (AP-PP, OP-DP); deltoid muscle thickness (at 3, 5 and 7 cm); distance of AXN and PCHA from the mid-acromial point in anatomical and 60 degrees abducted positions of arm. The data was tabulated, graphically represented and statistically analyzed.

### Risk assessment

Two investigators measured each and every variable individually and recorded their own observations. Both digital vernier calipers and anthropometric tape or thread were used for the measurements and the difference was assessed. To tackle the inter-observer bias, after completion of measurements of every specimen, both the investigators disclosed and discussed upon their findings and in case of more than 5% of difference in their measurements, the third investigator was called upon to reconduct the measurements or to comment on the differences. To address the intra-observer variability, two readings of every variable were taken and then the average was calculated. To combat the environmental factors, measurements were taken at mornings, afternoons and the evenings and again the average was calculated.

### Data collecting instruments

Anthropometry instruments including digital Vernier calipers, anthropometric tape, dissection instruments, specimen marking pins, dermato-graphic pencils, plastic ruler, and stationary items were used for data collection.

### Data analysis procedure

Data were analyzed using Microsoft Excel (Excel 2016, Microsoft Corp., Redmond, WA, USA) and mean, standard deviation was calculated. P value was estimated. The t-test was used to compare the variables of interest according to sex, and P value less than 0.05 was considered statistically significant. The present study was conducted in accordance with the principles of the Declaration of Helsinki.

## Results

The project was divided into an initial pilot period, followed by a continuation period. The objective of the pilot phase of 08 months (from 1^st^ October 2023 till 30^th^ May 2023) was to dissect the available six cadavers and the data arising out of these 6 cadavers i.e.12 specimens is presented in this article. The anthropometric data of the total 12 specimens was compiled in an Excel sheet, tabulated, graphically represented, and statistically analyzed as follows.

## Discussion

Ye-Gyung Kim et al. (Korea) and Nicholson et al. (USA) reported significant difference in the length of the acromion between males and females. Our results agree with those of these two studies [8,15]. Nakatani et al. (Japan) in their study observed that AXN is frequently positioned 5 cm below the mid-acromion point [16]. Our findings do not match this study. Nakajima et al. (Japan) recommended the periphery of deltoid tuberosity as the safest point for deltoid IM [9]. We found this site extremely close to the periosteum, with least thickness of deltoid muscle. Gurushantappa PK. (India) observed the distance of AXN from the midpoint of the postero-lateral border of the acromion process to be 7.46 cm +/- 0.99 cm [11]. In the present study, it was 8.19 cm. Patra A. et al. (India) reported that the mean distance of AXN from the anterior and posterior border of the acromion was 5.22 cm and 4.17 cm, respectively [10]. The present study’s mean length of AXN from OP was 8.19 +/- 0.616 cm. Compared with Nakajima et al.’s study, PCHA’s location differed from ours [9]. In Nakajima et al.’s and our study, the PCHA was 6.8 +/- 1 cm and 8.66 +/- 0.9 cm from mid-acromion respectively. Cheung K. et al. (USA) reported that the anterior branch of AXN moves 1.3 to 1.4 cms toward the acromion if the shoulder was abducted to 60 degrees [6]. In our study, AXN and PCHA moved 1.1 cm. Ye-Gyung Kim et al. (Korea) reported the mean distance of AXN and PCHA (0.5 cm below) from mid-acromial point to be 5.8 ± 1.0 cm and 6.3 ± 0.9 cm, respectively. The mean length of deltoid muscle was 16:1 ± 1:0 cm and the deltoid muscle was thickest at 7 cm. In our study, the mean deltoid length was 17.7 cm, 1 cm longer. The deltoid muscle was thickest at 7 cm. AXN and PCHA were at 8.19 +/- 0.61 and 8.66 +/- 0.96 cm, respectively.

International immunization advisory group namely ‘Centers for Disease Control and Prevention’ (CDC) (2021), ‘National Immunization Technical Advisory Groups’ (NITAGs) Ireland (2020) and New Zealand (2020) recommends the central and thickest portion, approximately 2 inches (5.08 cm) below the mid-acromion to be the safest site for deltoid IM [15]. As per our study findings, this site is the most dangerous site. According to the ‘Handbook of Safe Injection Practices’, released by the ‘National Centre for Disease Control’ (NCDC), Government of India, a site 1 to 2 inches (4.4 cm) below the acromion process is the safest site [16]. Our study observation contradicts this recommendation. United States Department of Health and Human Sciences Centres for Disease Control and Prevention (2017) reports that the safe site for deltoid IM is about 2-3 fingerbreadths below acromion process and above the armpit level of upper arm [17]. Our study observations contradicts these recommendations, the reason behind may be the racial differences in the deltoid muscle morphometry.

### Limitations of the study

The limited number of cadavers is the first limitations because of which we conducted a pilot study in the first half of the project. The movement of axillary nerve and posterior circumflex humeral artery towards the mid-acromial point is best observed in a living person, instead of cadavers for which an ultrasonographic study is recommended.

## Conclusions

The deltoid IM must be given at or below the level of midpoint of the deltoid muscle, but never in the upper half of deltoid muscle. We propose a site that lies 4 fingerbreadths/9 cm below the mid-acromion point as the safest site to avoid injury to the AXN, and PHCA. We propose to ask the vaccine recipient to place their hand on their hip thereby abducting the shoulder to 60 degrees and giving the deltoid IM into the mid-point of the muscle. Linear regression coefficients formulated are y= 1.132 x + 10.74. [x = length of acromion (AP-PP), y = length of deltoid (OP-DP)] and y= 0.0019 x + 8.16. [x = length of deltoid (OP-DP), y = length of acromion (AP-PP)].

## Data Availability

All data produced in the present work are contained in the manuscript

## Tables with titles and legends

**Table 1.** Measurements of reference line measurements (unit: centimeter) Table 1 shows that he length of acromial border (AP-PP) and of deltoid muscle (OP-DP) was higher in males compared to that in females.

| Measurements | Total<br>(mean+/-SD) | Males<br>(mean+/-SD) | Females<br>(mean+/-SD) | P value |
| --- | --- | --- | --- | --- |
| Length of acromial border (AP-PP) | 6.25+/- 0.602 | 6.35 +/- 0.620 | 5.8 +/- 0.141 | 0.017 (S) |
| Length of deltoid muscle (OP-DP) | 17.715 +/- 1.089 | 17.798 +/- 1.184 | 17.3+/- 0.155 | 0.115 (NS) |

SD: Standard deviation; AP: the anterior point of the acromion; PP: the posterior point of the acromion; OP: the origin points of the deltoid muscle; DP: the lowest point of the deltoid insertion; S: significant; NS: not significant.

**Table 2.** Measurements of the thickness of the deltoid muscle (unit: cm) Table 2 displays that the thickness of deltoid muscle was highest at 7 cm from the origin point (OP) than at 5 cm and 3 cm.

| Measurements | 3 cm | 5 cm<br>Mean (range) | 7 cm |
| --- | --- | --- | --- |
| Deltoid muscle thickness | 1.079 (0.5-1.78) | 1.599 (1-2.96) | 1.815 (1.2-2.5) |

SD: standard deviation

**Table 3.** Location of the main branches of the axillary nerve and the PCHA in relation to the OP (unit: centimeter) Table 3 shows that the axillary nerve lies at a mean distance of 8.19 cm (7.5 – 10 cm) from OP in both males and females. The PCHA lies at a mean distance of 8.25 cm and 8.69 cm from OP in males and females, respectively. PCHA pass 0.5 cm below the axillary nerve on an average.

| Measurements | Total<br>(mean+/-<br>SD) | Males<br>(mean+/-<br>SD) | Females<br>(mean+/-<br>SD) | Minimum | Maximum | P value |
| --- | --- | --- | --- | --- | --- | --- |
| Axillary nerve | 8.19+/-<br>0.616 | 8.19+/-<br>0.681 | 8.19+/-0.0 | 7.5 | 10 | 0.115<br>(NS) |
| Posterior<br>Circumflex<br>Humeral artery | 8.66 +/-<br>0.968 | 8.25 +/-<br>1.042 | 8.69 +/- 0.0 | 7.5 | 11 | 0.059<br>(NS) |

SD: Standard deviation; OP: the origin points of the deltoid muscle; OP: the origin points of the deltoid muscle; S: significant; NS: not significant.

**Table 4.** Location of the main branches of the axillary nerve and the PCHA in relation to the OP in 600 abduction (unit: centimeter). As per the table 4, the axillary nerve lies at a mean distance of 7.09 cm (6.4 – 8.9 cm) from OP in both males and females. The PCHA lies at a mean distance of 7.59 cm and 7.58 cm from OP in males and females, respectively. AXN and PCHA moved 1.1 cm towards the acromion in the abducted position.

| Measurements | Total<br>(mean+/-<br>SD) | Males<br>(mean+/-<br>SD) | Females<br>(mean+/-<br>SD) | Minimum | Maximum | P value |
| --- | --- | --- | --- | --- | --- | --- |
| Axillary nerve | 7.09 +/-<br>0.616 | 7.09 +/-<br>0.681 | 7.09 +/- 0.0 | 6.4 | 8.9 | 0.0487<br>(NS) |
| Posterior<br>Circumflex<br>Humeral artery | 7.56 +/-<br>0.968 | 7.59 +/-<br>1.042 | 7.58 +/- 0.0 | 6.4 | 9.9 | 0.059<br>(NS) |

SD: Standard deviation; OP: the origin points of the deltoid muscle; OP: the origin points of the deltoid muscle; S: significant; NS: not significant.

**Table 5.** Branching pattern of axillary nerve. Table 5 shows that the most common pattern of branching of axillary nerve was the first and the third ones. The figures 2-5 display the different branching patterns of axillary nerves.

| Type | Description | Number of<br>specimens | Percentage of<br>occurrence (%) |
| --- | --- | --- | --- |
| 1 | AXN pass above PCHA and gives 2 branches. | 04 | 33.3 |
| 2 | AXN pass above PCHA and gives more than 2 branches. | 03 | 25 |
| 3 | AXN pass below PCHA and divides into 2 branches. | 04 | 33.3 |
| 4 | AXN cross the PCHA from above to below. | 01 | 8.3 |

AXN: Axillary nerve; PCHA: Posterior circumflex humeral artery

**Figure 2:**
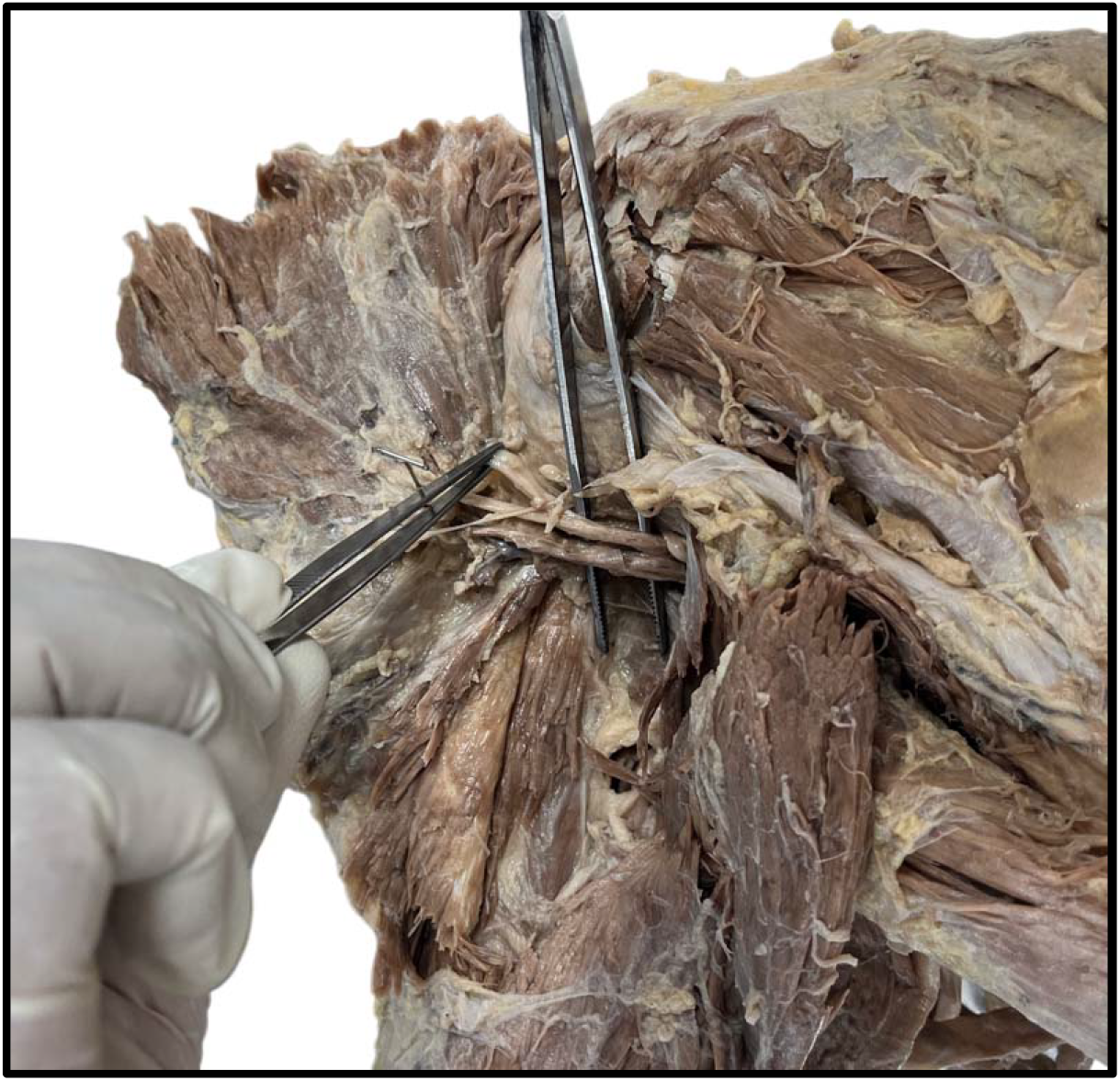
Specimen showing the Type −1 branching pattern of Axillary nerve.

**Figure 3:**
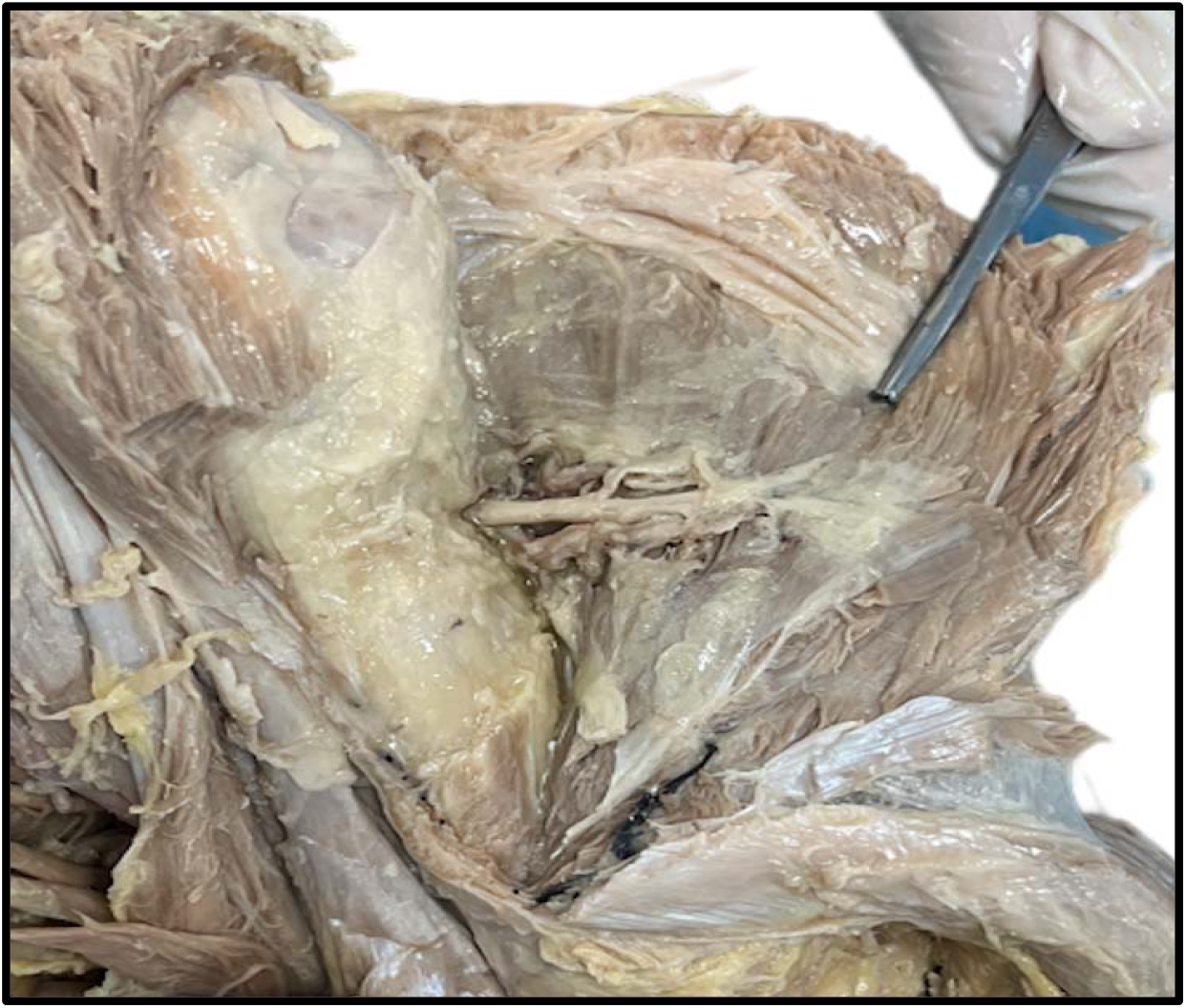
Specimen showing the Type −2 branching pattern of Axillary nerve.

**Figure 4:**
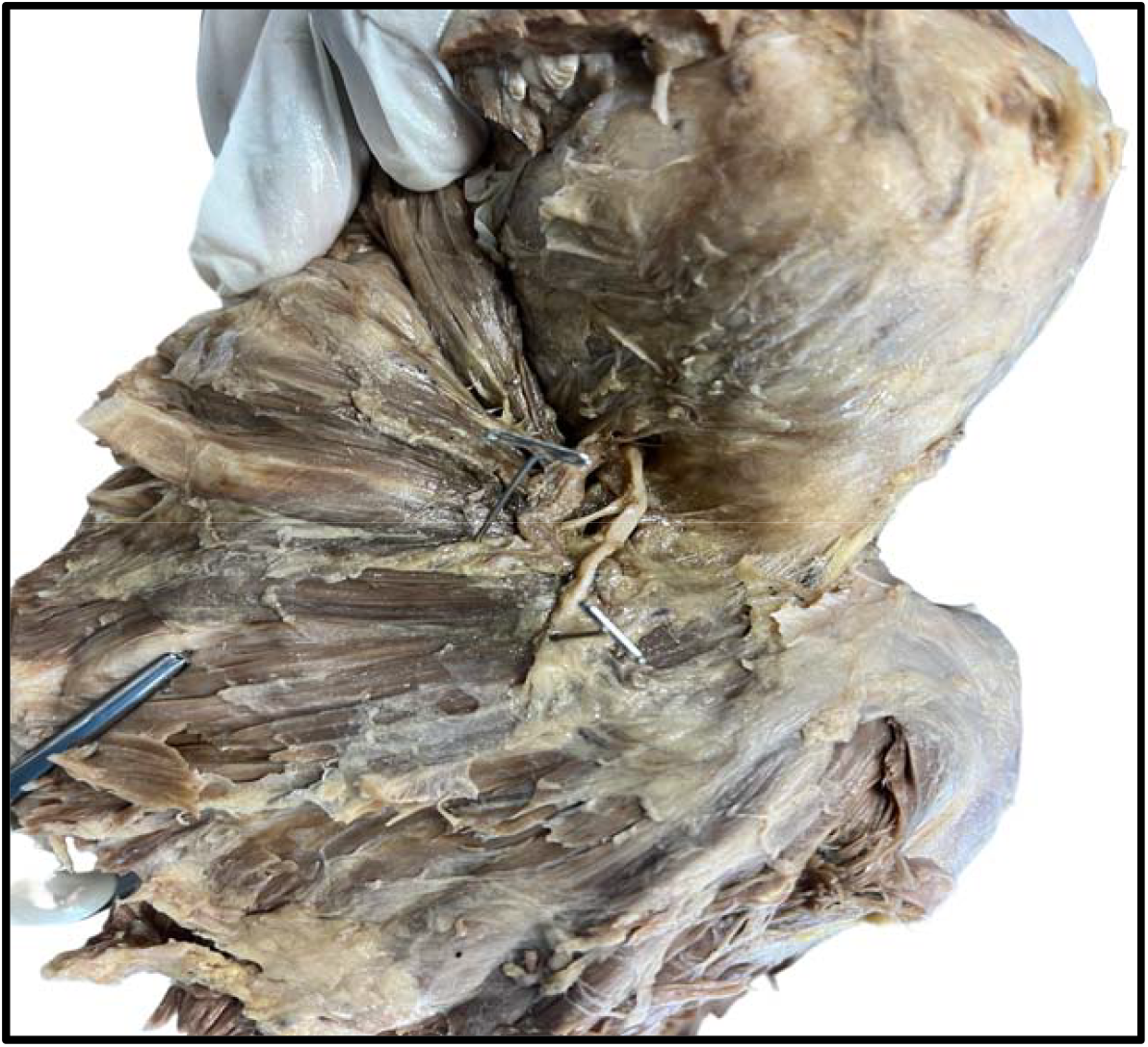
Specimen showing the Type −3 branching pattern of Axillary nerve.

**Figure 5:**
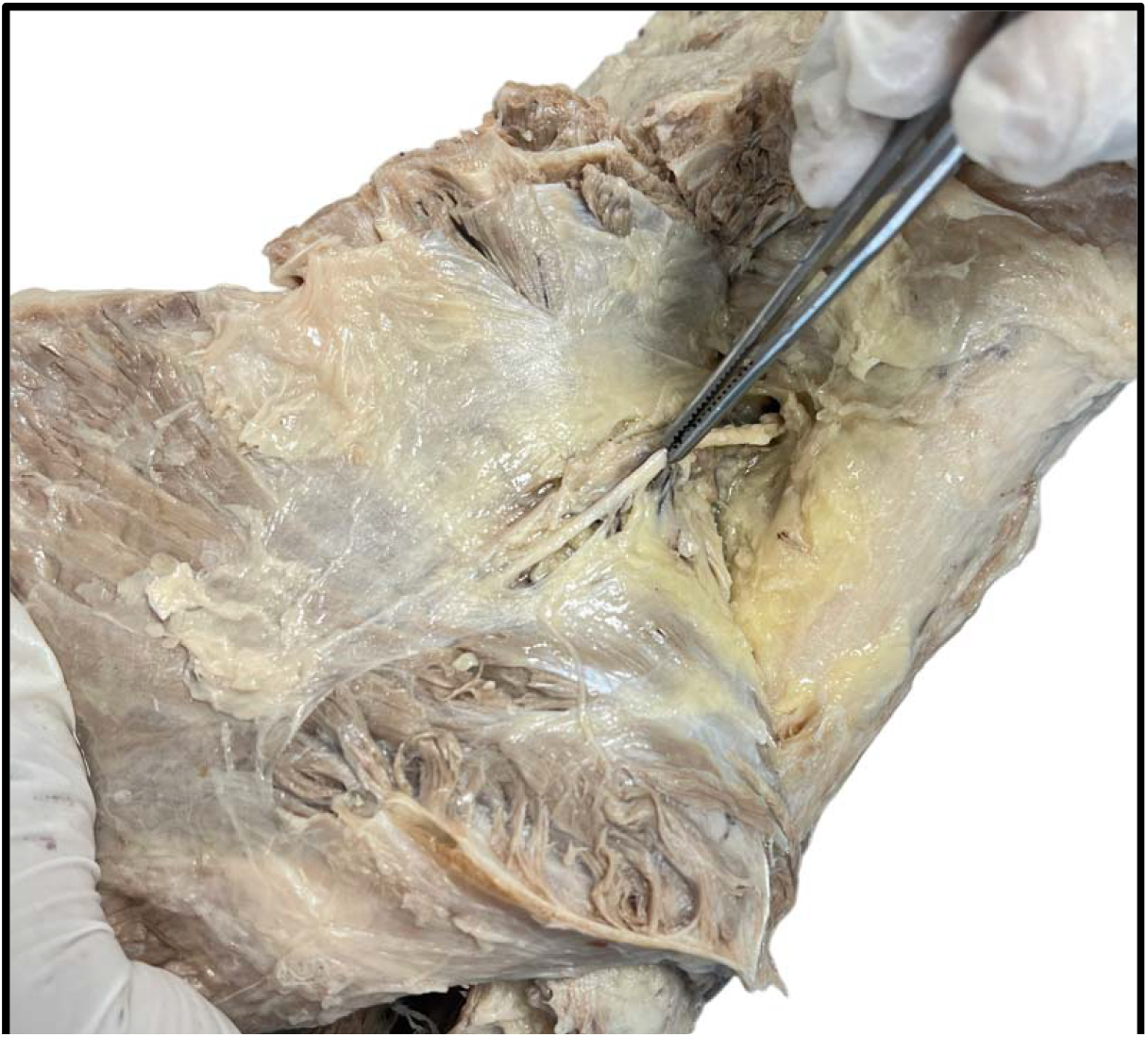
Specimen showing the Type −4 branching pattern of Axillary nerve.

